# Lysophosphatidic acid and lysophosphatidylcholine levels in serum samples of patients with major depressive disorder

**DOI:** 10.1101/2020.10.04.20206490

**Authors:** Sumaia Bari, Sharmin Sultana, Sohel Daria, Maliha Afrin Proma, Md. Rabiul Islam, Md. Ahsanul Haque

## Abstract

Major depressive disorder (MDD) is a heterogeneous condition featured with a continuous low mood, feeling of sadness, lack of interest to perform daily activities. Many factors including genetic, physiological, biological, social, and environmental are thought to be connected with the pathophysiology of depression. Several previous studies failed to identify the favorable biomarkers for MDD. Lysophosphatidic acid (LPA) and lysophosphatidylcholine (LPC)showed important roles in the regulation of emotion among experimental animals. The current study aimed to measure the serum levels of LPA and LPC in MDD patients and healthy controls (HCs) to explore their roles and relationship with depression. This case-control study enrolled 53 MDD patients and 50 healthy controls (HCs). The patients were recruited from the department of psychiatry, Bangabandhu Sheikh Mujib Medical University whereas the controls was from different locations of Dhaka city. Both the cases and controls were strictly matched by gender, age, and body mass index. A qualified psychiatrist diagnosed patients and evaluated controls based on the diagnostic and statistical manual of mental disorders, 5th edition. The severity of depression in MDD patients was measured by using the Hamilton depression rating scale (Ham-D). Enzyme-linked immunosorbent assay kits were used to measure serum levels LPA and LPC. We found no alterations of these parameters in serum levels of MDD patients compared to HCs. A significant positive correlation was found between serum LPA and LPC levels in MDD patients. Moreover, the present study showed no significant associations between target markers and either diagnosis of depression or Ham-D scores, or management of depression. The present study suggests that LPA and LPC levels probably would not serve as potential biomarkers of MDD. Thus, further studies with large and more homogeneous populations are recommended to explore the exact relationship between targeted serum lipids and major depression.

## 1. Introduction

Major depressive disorder (MDD) is a feeble and heterogeneous disease characterized by low mood, shortened interests, fatigue, concentration, ruined cognitive function, and developmental symptoms including oppressed sleep or appetite (Otte et al., 2016; Walther et al., 2018). MDD has become immensely dominant, enervating, and also a worthy illness in the present time (Kessler et al., 2003; Birnbaum et al., 2010; Judd et al., 2000). According to world health organization (WHO), MDD will become the second leading cause of disability throughout the world by 2020 because depression is one of the momentous disabling public health problems with the elevated rates of the outbreak (Kessler et al., 2003; Bilsker et al., 2006). Identification of a single causal factor for depression is difficult because the depressive disorder is a complex psychiatric condition where genetic variants, cytokine and neuroendocrine alterations, and environmental issues are involved (Belmmaker and Agam, 2008; Klibansky et al., 1957; Anjum et al., 2020; Munder et al., 1965; Nishuty et al., 2019; Heim et al., 2008).

There is no established mechanism or pathophysiology which can explain all the facts of MDD (Otte et al., 2016). The monoamine theory describes that the insufficient activity of monoamine-neuro-transmitters such as nor-adrenaline, dopamine, serotonin, etc. is the primary and major cause of depression (Islam et al., 2018). One of the researches suggested that decreased serotonergic neurotransmission can generate depression (Delgado and Moreno, 2000). A correlation between depression risk and polymorphisms in the 5-HTTLPR gene is found (Goldman et al., 2010). The correlation codes for serotonin receptors and suggests a link between monoamine neurotransmitters and MDD (Liu et al., 2018). Size reduction of the locus coeruleus, activity reduction of tyrosine hydroxylase enzyme, intensity enhancement of alpha-2 adrenergic receptor, and testimony from rat models recommend adrenergic neurotransmission reduction (Savitz and Drevets, 2013). A large number of researches have suggested hundreds of presumed biomarkers for the diagnosis, treatment, and prognosis of major depression. However, it is not yet discovered how the biologic information regarding biomarkers can be used to increase diagnosis, treatment, and prognosis. A potential target is provided by the biomarkers for determining prognosticators of response to MDD (Strawbridge et al., 2017). As MDD is highly comorbid, the neurobiological base for MDD is needed to be completely understood so that treatments for this disorder can always remain available and efficient (Müller et al., 2015).

It was found out by the researchers that malfunctions in neuronal proteins and peptide activities are one of the major causes of MDD (Duman, 2009; Emon et al., 2020; Ali et al., 2020). The lipidome which is also termed as the complete lipid profile of an organism-has a central part in most aspects of cell biology (Sud et al., 2007). The human brain mainly consists of around 60-70% lipids by dry weight (Svennerholm et al., 1994). Lipids that function structurally, mainly compose cell membranes and may be passively coordinated by several protein factors, whereas lipids function as signaling lipids through receptors in an extracellular fashion to trigger downstream pathways (Rossy et al., 2014). As lipids function as signaling medium they can easily promote depressive disorders and anxiety disorders (Müller et al., 2015). Lysophosphatidic acid (LPA) is one of the most essential reactive lipid species (Yung et al., 2015). LPA is synthesized and metabolized in a large number of metabolic pathways. Among all the pathways enzymatic action of autotaxin is the most crucial pathway (Perrakis and Moolenaar, 2014). The second important pathway is to obtain LPA from membrane phospholipids through the actions of phospholipase enzymes (Aoki et al., 2008). There are many additional pathways for producing LPA, particularly with the help of acyl-transferase enzymes (Pages et al., 2001). MDD is one of the most conventional psychiatric disorders, and the social burden of it is extremely huge (Ferrari et al., 2013). However, it is crucial to establish a biochemical maker regarding clinical purposes (Smith et al., 2013). LPA is identified as a potent bioactive lipid mediator with various biological characteristics. LPA plays very essential parts in the central nervous system. Biological importance and roles of LPA have been revealed by those studies (Aikawa et al., 2015). Due to the LPA receptor’s alteration, emotional behavior changes dramatically. An interesting matter is that LPA1-receptor knockout mice were reported to show abnormalities in their emotional behavior (Harrison et al., 2003). As blood brain-barrier protects the brain, intra-cerebro-ventricular injection of LPA was given to rats and mice, respectively, that has been shown to cause emotional alterations in rats (Castilla-Ortega et al., 2014) and mice (Yamada et al., 2014). Based on the above findings, LPA is one of the essential regulators of emotional behavior in the animal models, and thus possibly it may function in the pathophysiology of MDD.

Lysophosphatidylcholine (LPC) is also a kind of lipoprotein that may comply with a significant role in the neuronal mechanisms underlying the pathophysiology of several psychiatric disorders (Law et al., 2019). LPC is a water-soluble lipoprotein, amphiphilic molecule and it accumulates in micelles. The intentness of micelle relays on the chain length of the fatty acid molecule (Nakanaga et al., 2010). Though LPC is a lysosomal approach involved in central roles in most cases of cell biology the destabilization of the lysosomal process is troublesome for organelle and living cells (Hu et al., 2007). LPC is one of the leading phospholipids elements of oxidized low-density lipoprotein (Ox-LDL) also known as lysolecithins and produced from the biochemical transformation of phosphatidylcholine (PC) by phospholipase A2 enzyme present in cell membranes, Ox-LDL, etc. (Gauster et al., 2005). The promptness of diverse signaling pathways which are attached in production of oxidative stress and inflammatory responses by LPC because membrane formation and trafficking structured by amphipathic lipids including LPC and the signaling source is triggered through G protein-coupled receptor G2A and toll-like receptors (Walther et al., 2018; Law et al., 2019). Reports and also the meta-analyses research the activity of single lipids in the case of MDD suggest that polyunsaturated fatty acids could involve in the generation of MDD. Though lipids are sharply interconnected and hierarchically organized that’s why the mechanism of lipidome in MDD is indispensable (Walther et al., 2018). A report indicated that LPC deed as an effective biomarker for the pathogenesis of depression (Xiong et al., 2016).

From the above findings, it was observed that LPA and LPChave impacts on brain functioning and emotional behaviors are altered by them. For identifying the role of LPA and LPC in depression, a large number of researches have been implicated but its role in depressive illness is not yet clear. Different works of literature have different opinions regarding the association of LPA and LPC with major depression (Itagaki et al., 2019; Kim et al., 2018; Liu et al., 2016). Therefore, the present study aimed to analyze serum LPA and LPC levels in MDD patients and their corresponding controls to identify treatment specific biomarkers which may help to suggest better management of MDD either by medication or psychotherapy.

## 2. Methods

### 2.1 Study population

The present study included 53 drug-naïve MDD patients age ranging from 18 to 60 years and 50 healthy controls (HCs) matched by age, gender, and body mass index (BMI) during September 2019 to July 2020. The patient enrollment was completed from Bangabandhu Sheikh Mujib Medical University (BSMMU), Dhaka, Bangladesh. And, HCs were from diverse locations of Dhaka city. A specialized psychiatrist diagnosed and evaluated both the cases and controls according to the diagnostic and statistical manual of mental disorders, 5th edition (DSM-5). The coexistence of other complications was observed through a detailed neurological and physical screening. The severity scores of depression were measured using the Hamilton depression rating (Ham-D) scale. The subjects with no previous history of cardiovascular disease, epilepsy, hypertension, liver or kidney failure and had not been treated with any medication that could potentially alter the concentration of serum LPA and LPC levels were excluded from this study. Patients who were suffering from mental retardation and other comorbid psychiatric illness were also eliminated from the present study. Participants with substance abuse or dependency, severe organic conditions, excessive obesity, abnormal BMI, and the presence of infectious diseases were also eliminated. Socio-demographic data and different biophysical characteristics were recorded by using a structured-predesigned questionnaire.

### 2.2 Blood sample collectionand processing

A professional phlebotomist collected blood samples (5mL) of study participants from the cephalic vein by using a plastic syringe fitted with a stainless-steel needle. The collected blood samples were placed into falcon tubes and were allowed to clot for an hour at room temperature without any agitation. The serum samples were extracted from the collected blood samples by centrifugation at 1000 x g for 15 minutes at room temperature. The separated serum samples were immediately stored at −80□C until being used for further analysis.

### 2.3 Quantification of serum LPA and LPC

Commercially available enzyme-linked immunosorbent assay (ELISA) kits (Abbexa Ltd., UK) were used to measure the serum LPA and LPC levels according to the manufacturer’s instructions. The kits were based on competitive binding ELISA technology where antibodies specific to LPA and LPC were pre-coated onto the 96 well-plates. Briefly, 50μL of standard solution and serum sample were placed to the appropriate wells of the 96-well microtiter plate then the plates were gently shaken to mix the contents thoroughly. Then 50μL of detection reagent A added to each well and gently shaken the plates for proper mixing. The plates were then sealed and incubated at 37□C temperature for 1 hour. Then, the liquid in the wells was discarded by removing the cover and washed the plates properly using wash buffer. Then, 100μL of detection reagent B was dispensed into each well with gentle mixing and sealed the plates for 30 min incubation at 37°C. After repeating the washing process, 90μL tetramethylbenzidine (TMB) substrate was added into each well and the plates were again incubated for 15 min at 37°C. Finally, the reaction was stopped by adding 50μL of stop solution, and the absorbances were measured immediately at450nm. The serum LPA and LPC levels were calculated and presented as μmol/L. There was no cross-reactivity with other neurotrophic mediators and the coefficient of variance (CV) for intra-assay and inter-assay was<10% and <12%, respectively. The assay sensitivity for serum LPA and LPC were<52.7 ng/mL and <92.4 ng/mL, respectively.

### 2.4 Statistical analysis

All the collected data were checked and verified carefully to detect errors and rectified those accordingly. Then the coding, classification, and tabulation of data were performed. The independent sample t-test and Fisher’s exact test were performed for the continuous variables and categorical variables, respectively. Descriptive statistics were computed for the socio-demographic characteristics of patients. In this research, we used the box plot and scatter plot graphs to showcase the present study findings. The statistical package for social sciences (SPSS), version 25.0 was used for analyzing the data (Armonk, NY: IBM Corp.). The results were considered statistically significant where p-values were less than 0.05.

## 3. Results

The descriptive information about the study participants was presented in Table 1 and we observed that both the cases and controls were similar in terms of the stated parameters. The present study observed that most of the MDD patients were females and their BMI was within the normal range. Non-smoker population with medium economic impression was predominant in the present study. Following the past studies, the present study observed most of the MDD patients were in their third and fourth decades of life. Table 2 and Figure 1 demonstrated the clinical and laboratory findings of the study populations. Serum LPA and LPC levels did not show any significant changes in MDD patients compared to HCs (p> 0.05). Moreover, serum LPA and LPC concentrations did not correlate with age, gender, and BMI in the patient group. Figure 2 displays the correlations between LPA or LPC levels and the severity of MDD for both genders. Significant positive correlations were observed between serum LPA and LPC levels in all MDD patients (r = 739; p < 0.001), male MDD patients (r = 937; p < 0.001), and female MDD patients (r = 629; p < 0.001). But no significant correlation was obtained between LPA or LPC levels and Ham-D scores.

**Table 1.**
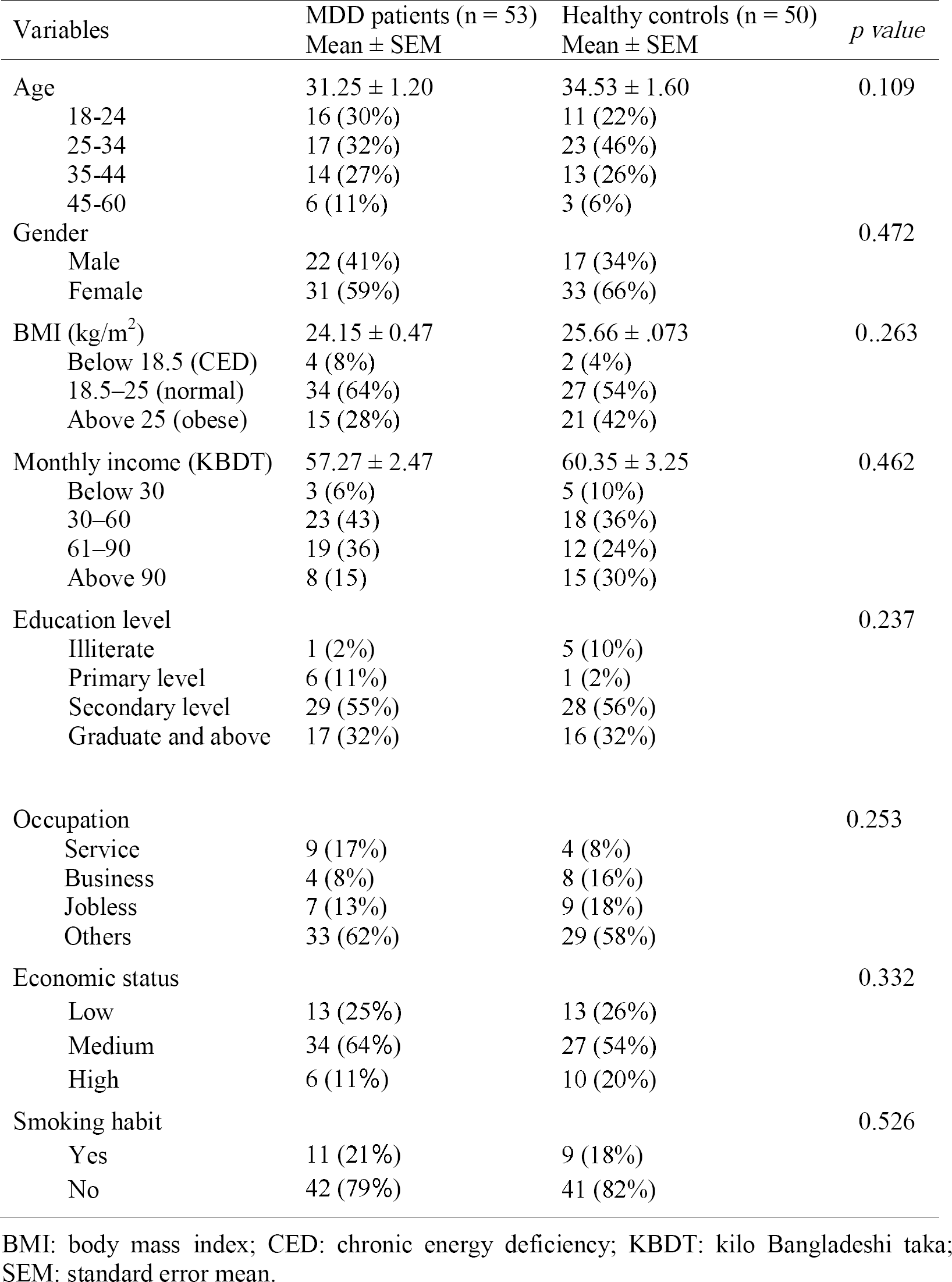
Socio-demographic characteristics of the study population.

**Table 2.** Clinical characteristics and laboratory findings of the study population.

| Variables | MDD patients (n = 53)<br>Mean ± SEM | Healthy controls (n = 50)<br>Mean ± SEM | *p value* |
| --- | --- | --- | --- |
| Ham-D score | 15.90 ± 0.57 | 3.61 ± 0.44 | <0.001 |
| Serum LPA (μmol/L) | 0.59 ± 0.09 | 0.58 ± 0.08 | 0.543 |
| Male | 0.58 ± 0.15 | 0.59 ± 0.10 | 0.991 |
| Female | 0.60 ± 0.12 | 0.70 ± 0.11 | 0.552 |
| Serum LPC (μmol/L) | 161.30 ± 22.96 | 170.87 ± 22.16 | 0.790 |
| Male | 192.10 ± 42.39 | 149.72 ± 28.57 | 0.413 |
| Female | 139.44 ± 25.21 | 176.79 ± 21.82 | 0.264 |

Ham-D: Hamilton depression rating scale; LPA: Lysophosphatidic acid; LPC: Lysophosphatidylcholine; SEM: Standard error mean.

**Fig. 1.**
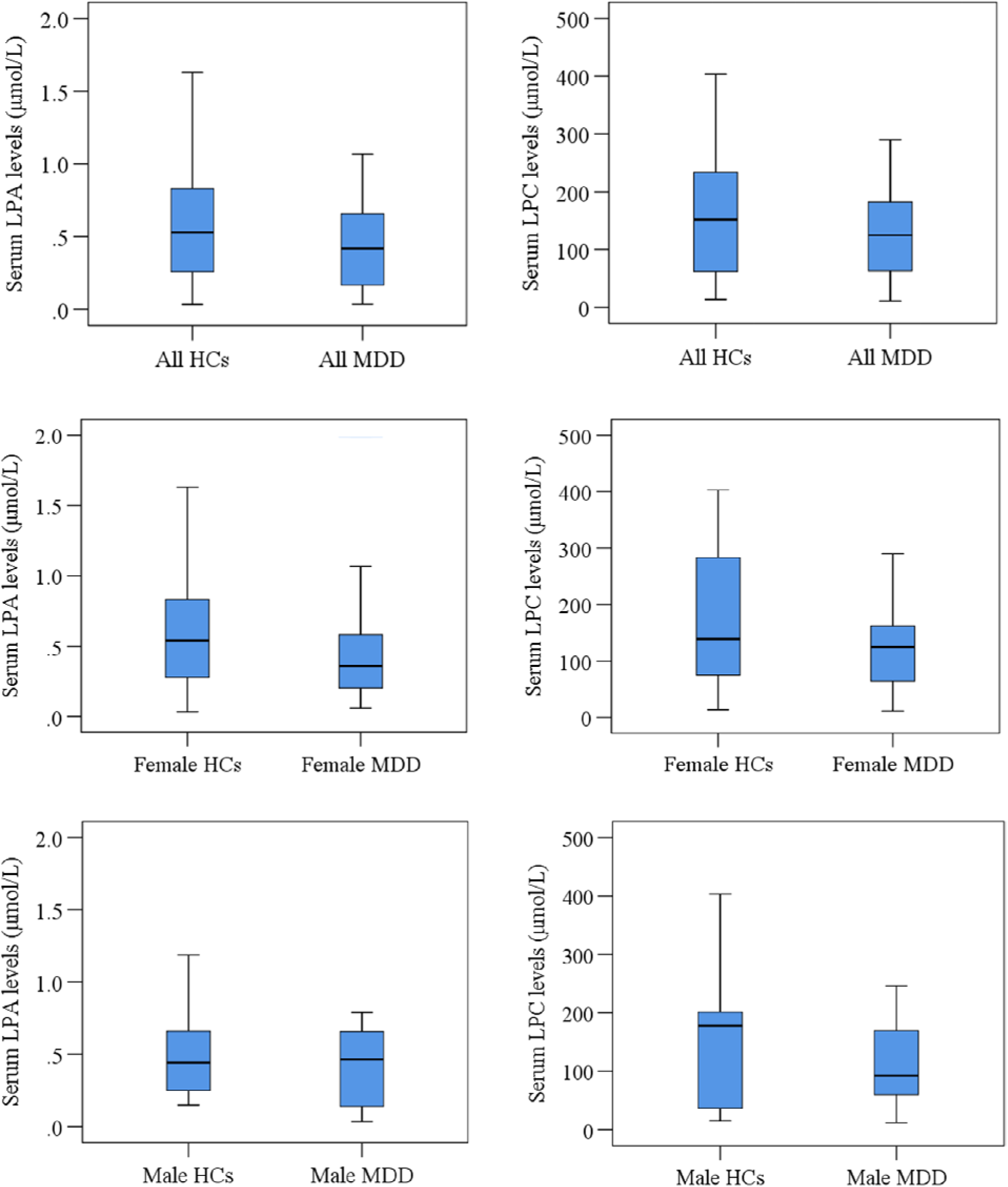
Changes of serum lysophosphatidic acid (LPA) and lysophosphatidylcholine (LPC) among the study population. HCs: healthy controls; MDD: major depressive disorder.

**Fig. 2.**
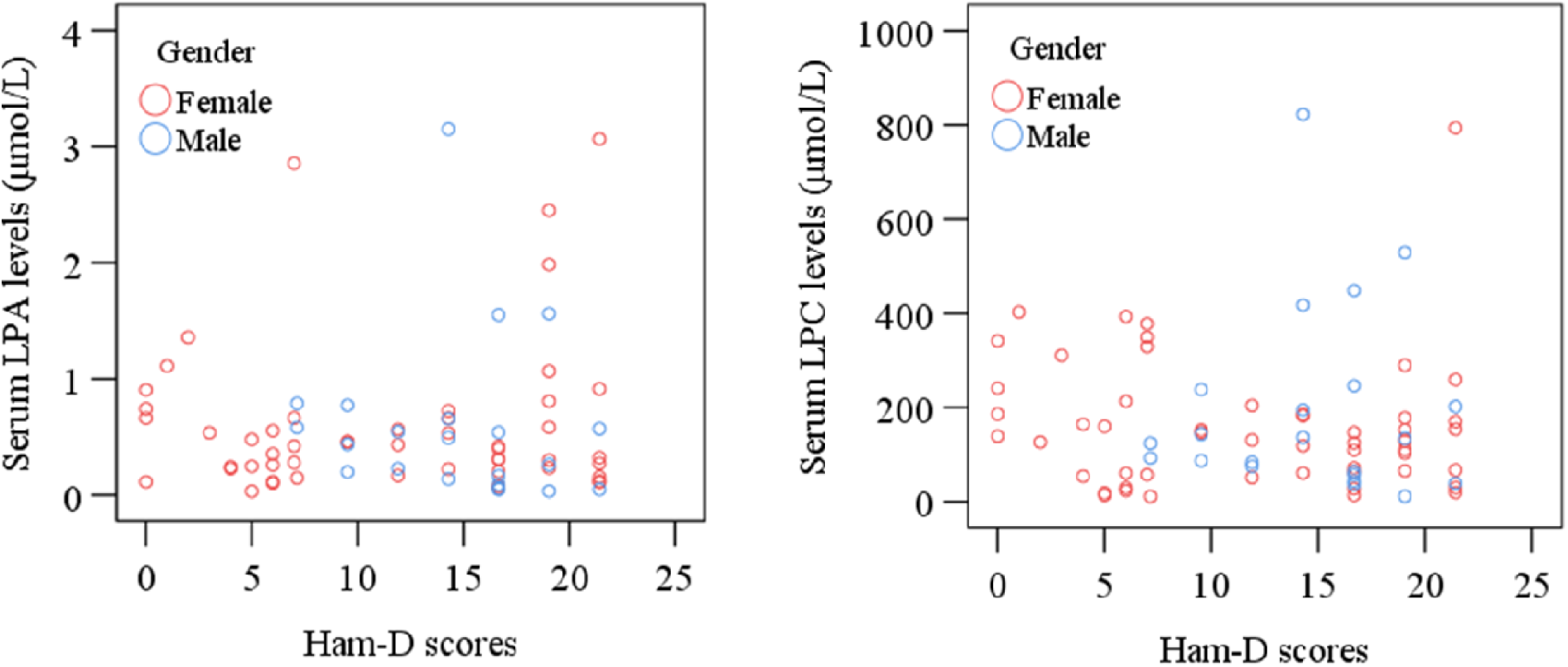
Correlations of serum lysophosphatidic acid (LPA) and lysophosphatidylcholine (LPC) with Ham-D scores in MDD patients were assessed by Pearson’s correlation coefficient.

## 4. Discussion

To the best of our knowledge, this is the first report concerning serum LPA and LPC in MDD patients from Bangladesh. In the present study, we evaluated whether LPA or LPC levels could serve as an early risk assessment marker for MDD. The diagnosis of depression is mainly depending on the structured clinical interview, and a reliable marker is not yet established for clinicians (Smith et al., 2013). Moreover, the serum LPA levels in MDD patients were reported significantly decreased compared to HCs (Itagaki et al., 2019; Kim et al., 2018) Liu et al. reported that plasma LPC levels remarkably elevated in MDD patients and showed a significant positive correlation with the severity of depression (Liu et al., 2016; Liu et al., 2015) Kim et al. also observed that serum LPC levels were decreased in drug-treated or drug-naïve MDD patients compared to HCs (Kim et al., 2018). An animal model study reported that serum LPC levels were significantly lower in depression than that of controls whereas another study contrasting the results by showing elevated plasma levels of LPC among the depressed rodents (Xiong et al., 2016; Chen et al., 2014). Thus, we verified the hypothesis that LPA and LPC levels in serum are connected with the early risk assessment or diagnosis of MDD. Despite the logic of our hypothesis, we observed that no significant alterations of serum LPA and LPC levels were happened in MDD patients compared to HCs, and therefore these lipids are unlikely to serve as early risk assessment markers of MDD. The present study is consistent with the past findings where Gotoh et al. observed that LPA levels in cerebrospinal fluid (CSF) and plasma were not associated with MDD (Gotoh et al., 2019a; Gotoh et al., 2019b). LPA is produced from LPC with the enzyme autotoxin (ATX). Enzymatic production of LPC from LPA makes it suitable to enter the brain crossing the blood-brain barrier with the help of the G2A receptor of transport protein (Nakanaga et al., 2010; Tsuda et al., 2006). The present study results along with the previous report, we reasonably assume that LPA levels could not be termed as the applicant marker for MDD.

Finally, correlation coefficients were measured between serum LPA or LPC levels with patient characteristics and clinical severity scores of depressions. No significant relationships were achieved between either serum LPA or LPC levels and patient profile such as age, gender, BMI, education, occupation, economic impression, and smoking history. Also, we did not find any significant positive or negative correlation between serum LPA or LPC levels and severity of depression. The present study results suggest that serum LPA or LPC levels would not play a significant role as a biomarker for clinical diagnosis of major depression. The above findings are consistent with the previous study results (Liu et al., 2015).

This present study has a few limitations. We only enrolled patients from the first depressive episode, serum samples from MDD patients with pre-and post-treatment with antidepressant medication might show different results. This was a case-control study and the small sample size might also intervene with the findings. Therefore, further studies examining the correlation between LPA or LPC signaling system and antidepressant medication in MDD patients are recommended to discover the role of lipids on the pathophysiology of depression.

## 5. Conclusions

The present study concluded that serum LPA and LPC levels would not serve as risk assessment markers for MDD. Serum levels of these lipids did not show any changes in MDD patients compared to HCs and also failed to show any correlation with the severity of depression. As a result, the present study is suggesting the association between serum LPA or LPC levels and the pathophysiology of depression is not well established. Hence, the current study should be taken as preliminary and further researches are recommended to investigate the exact correlation between targeted glycerophospholipids and major depression.

## Data Availability

The data supporting this study will be available from the corresponding author upon reasonable request.

## Declarations

### Author contribution statement

SB and SS conceived the idea, designed the study, collected the data, prepared the initial draft of the manuscript. SD and MAP contributed to the development of the study design, collected the data, edited the manuscript. MRI and MAH conceived and designed the study, analyzed and interpreted the data, reviewed the manuscript for important intellectual content, and supervised the whole work. All the authors approved the final version of the manuscript for this submission.

### Ethical approval

The study was conducted according to the world medical association declaration of Helsinki. All study participants gave written consent to participate in this study. The study protocol was approved by the ethical review committee of the department of psychiatry, BSMMU, Dhaka, Bangladesh.

## Acknowledgments

We are thankful to the physicians and staff of the department of psychiatry, BSMMU, Dhaka, Bangladesh, for their support in this study. We are also thankful to the participants of this study for their cooperation and participation.

## Additional information

No additional information is available.

## References

Otte, C., Gold, S. M., Penninx, B. W., Pariante, C. M., Etkin, A., Fava, M., Schatzberg, A.F., 2016. Major depressive disorder. Nat. Rev. Dis. Prim. 2, 16065.

Walther, A., Cannistraci, C. V., Simons, K., Durán, C., Gerl, M. J., Wehrli, S., Kirschbaum, C., 2018. Lipidomics in major depressive disorder. Front. Psychiatr. 9.

Kessler, R. C., Berglund, P., Demler, O., Jin, R., Koretz, D., Merikangas, K. R., Wang, P. S.,2003. The epidemiology of major depressive disorder. JAMA. 289, 3095.

Birnbaum, H. G., Kessler, R. C., Kelley, D., Ben-Hamadi, R., Joish, V. N., Greenberg, P. E., 2010. Employer burden of mild, moderate, and severe major depressive disorder: mental health services utilization and costs, and work performance. Depress. Anxiet. 27, 78–89.

Judd, L.L., Akiskal, H.S., Zeller, P.J., Paulus, M., Leon, A.C., Maser, J.D., Keller, M.B., 2000. Psychosocial disability during the long-term course of unipolar major depressive disorder. Arch. Gen. Psychiatr. 57, 375.

Bilsker, D., Wiseman, S., Gilbert, M., 2006. Managing depression-related occupational disability: A pragmatic approach. Can. J. Psychiatr. 51, 76–83.

Belmaker, R.H., Agam, G., 2008. Major depressive disorder. N. Engl. J. Med. 358, 55–68.

Klibansky, C., Smorodinsky, I., de Vries, A., 1967. Action of lysolecithin on normal and leukemic human leukocytes. Med. Pharmacol. Exp.16, 297–304.

Anjum, S., SalahudinQusar, M.M.A., Shahriar, M., Islam, S.M.A., Bhuiyan, M.A., Islam, M.R., 2020. Altered serum interleukin-7 and interleukin-10 are associated with drug-free major depressive disorder. Ther. Adv. Psychopharmacol. 10, 204512532091665.

Munder, P.G., Ferber, E., Fischer, H., 1965. Untersuchungen uber die Abhangigkeit der cytolytischenWirkung des Lysolecithins von Membranenzymen [Studies on the dependence of the cytolytic effect of lysolecithin on cell membrane enzymes]. Z. Naturforsch. B. 20, 1048–1061.

Nishuty, N.L., Khandoker, M.M.H., Karmoker, J.R., Ferdous, S., Shahriar, M., SalahuddinQusar, M.M.A., Islam, M.S., Kadir, M.F., Islam, M.R., 2019. Evaluation of serum Interleukin-6 and C-reactive protein levels in drug-naïve major depressive disorder patients. Cureus 11, e3868.

Heim, C., Newport, D.J., Mletzko, T., Miller, A.H., Nemeroff, C.B., 2008. The link between childhood trauma and depression: insights from HPA axis studies in humans. Psychoneuroendocrin. 33, 693–710.

Islam, M.R., Islam, M.R., Ahmed, I., Moktadir, A.A., Nahar, Z., Islam, M.S., Hasnat, A. 2018. Elevated serum levels of malondialdehyde and cortisol are associated with major depressive disorder: A case-control study. SAGE Open Med. 6, 205031211877395.

Delgado, P.L., Moreno, F.A., 2000. Role of norepinephrine in depression. J. Clin. Psychiatr. 61 (Suppl 1), 5–12.

Goldman, N., Glei, D.A., Lin, Y.H., Weinstein, M., 2010. The serotonin transporter polymorphism (5-HTTLPR): allelic variation and links with depressive symptoms. Depress. Anxiet. 27, 260–269.

Liu, Y., Zhao, J., Guo, W., 2018. Emotional roles of mono-aminergic neurotransmitters in major depressive disorder and anxiety disorders. Front. Psychol. 9, 2201.

Savitz, J.B., Drevets, W.C., 2013. Neuroreceptor imaging in depression. Neurobiol. Dis. 52, 49–65.

Strawbridge, R., Young, A.H., Cleare, A.J., 2017. Biomarkers for depression: recent insights, current challenges and future prospects. Neuropsychiatr. Dis. Treat. 13, 1245–1262.

Müller, C.P., Reichel, M., Mühle, C., Rhein, C., Gulbins, E., Kornhuber, J., 2015. Brain membrane lipids in major depression and anxiety disorders. BiochimBiophys. Acta. 1851, 1052–1065.

Duman, R.S., 2009. Neuronal damage and protection in the pathophysiology and treatment of psychiatric illness: stress and depression. Dialogues Clin. Neurosci. 11, 239–255.

Emon, M.P.Z., Das, R., Nishuty, N.L., ShalahuddinQusar, M.M.A., Bhuiyan, M.A., Islam, M.R., 2020. Reduced serum BDNF levels are associated with the increased risk for developing MDD: a case-control study with or without antidepressant therapy. BMC. Res. Notes. 13, 83.

Ali, S., Nahar, Z., Rahman, M.R., Islam, S.M.A., Bhuiyan, M.A., Islam, M.R., 2020. Serum insulin-like growth factor-1 and relaxin-3 are linked with major depressive disorder. Asian. J. Psychiatr. 53, 102164.

Sud, M., Fahy, E., Cotter, D., Brown, A., Dennis, E. A., Glass, C. K., Subramaniam, S., 2007. LMSD: LIPID MAPS structure database. Nucleic Acids Res. 35(Database), D527–D532.

Svennerholm, L., Boström, K., Jungbjer B., Olsson, L., 1994. Membrane lipids of adult human brain: lipid composition of frontal and temporal lobe in subjects of age 20 to 100 years. J. Neurochem. 63, 1802–1811.

Rossy, J., Ma, Y., Gaus, K., 2014. The organisation of the cell membrane: do proteins rule lipids? Curr. Opin. Chem. Biol. 20, 54–59.

Yung, Y.C., Stoddard, N.C., Mirendil, H., Chun, J., 2015. Lysophosphatidic Acid signaling in the nervous system [published correction appears in Neuron. Neuron 85, 669–682.

Perrakis, A., Moolenaar, W.H., 2014. Autotaxin: structure-function and signaling. J. Lipid. Res. 55, 1010–1018.

Aoki, J., Inoue, A., Okudaira, S., 2008. Two pathways for lysophosphatidic acid production. Biochim. Biophys. Acta. 781, 513–518.

Pagès, C., Simon, M.F., Valet, P., Saulnier-Blache J.S., 2001. Lysophosphatidic acid synthesis and release. Prostaglandins Other Lipid Mediat. 64, 1–10.

Ferrari, A. J., Charlson, F. J., Norman, R. E., Flaxman, A. D., Patten, S. B., Vos, T., Whiteford, H. A., 2013. The epidemiological modelling of major depressive disorder: Application for the global burden of disease study 2010. PLoS ONE 8, e69637.

Smith, K.M., Renshaw, P.F., Bilello, J., 2013. The diagnosis of depression: current and emerging methods. Compr. Psychiatry. 54, 1–6.

Aikawa, S., Hashimoto, T., Kano, K., Aoki, J., 2015. Lysophosphatidic acid as a lipid mediator with multiple biological actions. J. Biochem. 157, 81–9.

Harrison, S. ., Reavill, C., Brown, G., Brown, J. ., Cluderay, J. ., Crook, B., Maycox, P., 2003. LPA1 receptor-deficient mice have phenotypic changes observed in psychiatric disease. Mol. Cell. Neurosci. 24, 1170–1179.

Castilla-Ortega, E., Escuredo, L., Bilbao, A., Pedraza, C., Orio, L., Estivill-Torrús, G., Pavón, F. J., 2014. 1-Oleoyl Lysophosphatidic Acid: A New Mediator of Emotional Behavior in Rats. PLoS ONE 9, e85348.

Yamada, M., Tsukagoshi, M., Hashimoto, T., Oka, J.-I., Saitoh, A., Yamada, M., 2014. Lysophosphatidic acid induces anxiety-like behavior via its receptors in mice. J. Neural Transm. 122, 487–494.

Law, S.H., Chan, M.L., Marathe, G.K., Parveen, F., Chen, C.H., Ke, L.Y., 2019. An Updated Review of Lysophosphatidylcholine Metabolism in Human Diseases. Int. J. Mol. Sci. 20, 1149.

Nakanaga, K., Hama, K., Aoki, J., 2010. Autotaxin—a LPA producing enzyme with diverse functions. J. Biochem. 148, 13–24.

Hu, J.S., Li, Y.B., Wang, J.W., Sun, L., Zhang, G.J., 2007. Mechanism of Lysophosphatidylcholine-Induced Lysosome Destabilization. J. Membr. Biol. 215, 27– 35.

Gauster, M., Rechberger, G., Sovic, A., Hörl, G., Steyrer, E., Sattler, W., Frank, S., 2005. Endothelial lipase releases saturated and unsaturated fatty acids of high density lipoprotein phosphatidylcholine. J. Lipid Res. 46, 1517–1525.

Xiong, Z., Yang, J., Huang, Y., Zhang, K., Bo, Y., Lu, X., Wu, C., 2016. Serum metabonomics study of anti-depressive effect of Xiao-Chai-Hu-Tang on rat model of chronic unpredictable mild stress. J. Chromatogr. B. Analyt. Technol. Biomed. Life Sci. 1029-1030, 28–35.

Itagaki, K., Takebayashi, M., Abe, H., Shibasaki, C., Kajitani, N., Okada-Tsuchioka, M., Yamawaki, S., 2019. Reduced serum and cerebrospinal fluid levels of autotaxin in major depressive disorder. Int. J. Neuropsychopharmacol. 22, 261–269.

Kim, E.Y., Lee, J.W., Lee, M.Y., Kim, S.H., Mok, H.J., Ha, K., Kim, K.P., 2018. Serum lipidomic analysis for the discovery of biomarkers for major depressive disorder in drug-free patients. Psychiatr. Res. 265, 174–182.

Liu, X., Li, J., Zheng, P., Zhao, X., Zhou, C., Hu, C., Xu, G., 2016. Plasma lipidomics reveals potential lipid markers of major depressive disorder. Anal. Bioanal. Chem. 408, 6497– 6507.

Liu, X., Zheng, P., Zhao, X., Zhang, Y., Hu, C., Li, J., Xu, G., 2015. Discovery and Validation of Plasma Biomarkers for Major Depressive Disorder Classification Based on Liquid Chromatography–Mass Spectrometry. J. Proteome Res. 14, 2322–2330.

Chen, S., Wei, C., Gao, P., Kong, H., Jia, Z., Hu, C., Xu, G., 2014. Effect of Allium macrostemon on a rat model of depression studied by using plasma lipid and acylcarnitine profiles from liquid chromatography/mass spectrometry. J. Pharm. Biomed. Anal. 89, 122–129.

Gotoh, L., Yamada, M., Hattori, K., Sasayama, D., Noda, T., Yoshida, S., Yamada, M., 2019a. Lysophosphatidic acid levels in cerebrospinal fluid and plasma samples in patients with major depressive disorder. Heliyon 5, e01699.

Gotoh, L., Yamada, M., Hattori, K., Sasayama, D., Noda, T., Yoshida, S., Yamada, M., 2019b. Levels of lysophosphatidic acid in cerebrospinal fluid and plasma of patients with schizophrenia. Psychiatry. Res. 273, 331–335.

Tsuda, S., Okudaira, S., Moriya-Ito, K., Shimamoto, C., Tanaka, M., Aoki, J., Kobayashi, T., 2006. Cyclic Phosphatidic Acid Is Produced by Autotaxin in Blood. J. Biol. Chem. 281, 26081–26088.

